# An assessment of the value of deep neural networks in genetic risk prediction for surgically relevant outcomes

**DOI:** 10.1101/2024.01.09.23297913

**Authors:** Mathias A Christensen, Arnór Sigurdsson, Alexander Bonde, Simon Rasmussen, Sisse R Ostrowski, Mads Nielsen, Martin Sillesen

## Abstract

**Introduction:** Postoperative complications affect up to 15% of surgical patients constituting a major part of the overall disease burden in a modern healthcare system. While several surgical risk calculators have been developed, none have so far been shown to decrease the associated mortality and morbidity. Combining deep neural networks and genomics with the already established clinical predictors may hold promise for improvement.

**Methods:** The UK Biobank was utilized to build linear and deep learning models for the prediction of surgery relevant outcomes. An initial GWAS for the relevant outcomes was initially conducted to select the Single Nucleotide Polymorphisms for inclusion in the models. Model performance was assessed with Receiver Operator Characteristics of the Area Under the Curve and optimum precision and recall. Feature importance was assessed with SHapley Additive exPlanations.

**Results:** Models were generated for atrial fibrillation, venous thromboembolism and pneumonia as genetics only, clinical features only and a combined model. For venous thromboembolism, the ROC-AUCs were 59.6% [59.0%-59.7%], 63.4% [63.2%-63.4%] and 66.1% [65.7%-66.1%] for the linear models and 60.0% [57.8%-61.8%], 63.2% [61.2%-65.0%] and 65.4% [63.6%-67.2%] for the deep learning SNP, clinical and combined models, respectively. For atrial fibrillation, the ROC-AUCs were 60.9% [60.6%-61.0%], 78.7% [78.7%-78.7%] and 80.1% [80.0%-80.1%] for the linear models and 59.9% [.6%-61.3%], 78.8% [77.8%-79.8%] and 79.4% [78.8%-80.5%] for the deep learning SNP, clinical and combined models, respectively. For pneumonia, the ROC-AUCs were 57.3% [56.5%-57.4%], 69.2% [69.1%-69.2%] and 70.5% [70.2%-70.6%] for the linear models and 55.5% [54.1%-56.9%], 69.7% [.5%-70.8%] and 69.9% [68.7%-71.0%] for the deep learning SNP, clinical and combined models, respectively.

**Conclusion:** In this report we presented linear and deep learning predictive models for surgery relevant outcomes. Overall, predictability was similar between linear and deep learning models and inclusion of genetics seemed to improve accuracy.

## INTRODUCTION

Worldwide, more than 310 million surgeries are performed each year, addressing an estimated 11% of the global burden of disease.(1, 2) While most surgical patients proceed to an uneventful recovery, current estimates indicate that roughly 4% die as a direct or indirect result of surgery, while up to 15% experience a postoperative complication (PC), prolonging hospital length-of-stay with consequential morbidity.(2)

While treatment advances following the implementation of approaches such as Enhanced Recovery after Surgery (ERAS) protocols have been well documented, the incidences of PCs have remained remarkably stable over the last decade.(3) As such, a stable subset of patients still experiences PCs, suggesting that this patient group could potentially benefit from a deviation from the current one-size-fits all approach deployed by most ERAS protocols and a move towards a precision medicine approach in the surgical setting.

To achieve this goal, risk predictions models are, however, needed for the identification of patients who will fail standard ERAS protocols.

To this end, many risk assessment tools have been fielded to identify at-risk patients including the regression-based American College of Surgeons National Surgical Quality Improvement Program (ACS-NSQIP) risk calculator as well as newer machine learning approaches investigating the value of random forests or deep neural networks (DNNs).(4, 5) These models are, however, limited by the fact that they only perform predictions on available clinical data, which may provide insights into parts of the driving factors of the patients risks only.

As such, recent data has suggested that genetic susceptibility could, in part, be a modifier of individual PCs risk, thus opening the potential for adding genetic data points to risk prediction models in order to improve model performance.(6, 7)

Genetic variations are increasingly being recognized as an important modality for various surgical adverse events including venous thromboembolisms, renal complications and cardiac arrythmias.(6, 8, 9) However, it is currently not clear to what degree genetic susceptibility contribute to the overall risk compared with other well-known clinical risk factors. Furthermore, as genetic susceptibility may include complex non-linear effects such as previously non-identified complex interactions between genes that possibly lie far from each other in the human genome, optimal modelling strategies remain unknown. As such, whether legacy risk prediction approaches such as the linear Polygenic Risk Scores (PGS), traditionally utilized to assess an overall genetic risk composition and weighted sum for the phenotype in question, could be inferior to a DNN approach, is currently unknown.(10) Using the clinical question of assessing whether DNNs can outperform a classic PGS approach for assessing genotype-associated risk of PCs, we target three high impact PCs with proven genetic susceptibility.(11) These include postoperative pneumonias, postoperative venous thromboembolisms (pVTEs) and postoperative atrial fibrillation (pAFLI). Furthermore, we investigate whether single nucleotide polymorphisms (SNPs) highlighted as driving the phenotype, differ between DNN and PGS approaches, thus potentially indicating that non-linear genotype-phenotype associations can be identified by the DNN approach. We hypothesize that DNNs will achieve superior predictive performance in predicting the genotype-associated risk of these PCs compared with a linear PGS, and that the DNN models will highlight a different subset of important SNPs compared with a linear PGS.

## METHODS

This study utilized genotype data from the United Kingdom biobank (UKB) consortium.(12) Access to the UKB data was approved by the consortium (Study ID #60861). Under Danish law, the study was exempt from ethical board approval due to the anonymized nature of the dataset.

We conducted a comparative study of different methodologies for genotyping risk prediction and Single Nucleotide Polymorphism (SNP)-identification in a general as well as a surgical, national cohort.

For the initial approach, we conducted standard GWAS-analyses without covariates on the chosen phenotypes with a high prevalence following surgery. Details for the GWAS are described below. These phenotypes included venous thromboembolisms (VTE), atrial fibrillation (AF) and bacterial pneumonia.

UKB has more than 500,000 individuals enrolled and consented across the United Kingdom of the age from 40 to 69. Patients were invited for participation through National Health service (NHS) registries and asked to fill surveys on basic demographic data, general lifestyle measures as well as medical history. Inclusion of all participants took place from 2006 to 2010.

### Identification of cohort

All patients with available genomic data in the UKB were initially included for analysis. Cases were identified depending on the phenotype in question. For AF, VTE and pneumonia, cases were defined using relevant *International Statistical Classification of Disease, 9^th^ revision* (ICD-9) and ICD-10 codes.

The phenotypes in question were identified with the ICD-9 and ICD-10 codes listed in supplementary table 1. The cohorts were split into training/validation and test sets. The training/validation set consisted of all non-surgical patients and a random sample of 80% of the surgical cohort. The test set consisted of the remaining 20% of the surgical cohort. Surgery was defined with the OPCS-4 codes listed in supplementary table 2. The post-surgical phenotypes were defined with the same ICD-codes as above registered up to 30 days after the given procedure. For AF, only first-time diagnoses were counted as post-surgery cases. For VTE and pneumonia, any diagnoses within 30 days were counted as cases, regardless of previous history.

For each outcome of interest (pAFLI, pVTE and pneumonia, both deep learning and linear models were created using three distinct input strategies (see below for model descriptions):

1. A genotype only model: using only the identified SNPs (see below) as input (SNP model)
2. A clinical data only model: using only clinical data as input (Clinical model)
3. A combined model: using both SNPs and clinical data as input (Combined model)

Input SNPs were the top 100 SNPs from the discovery GWAS for each phenotype of interest, with clinical data including demographics and comorbidities (supplementary table 3) and combined models including both genetic and clinical data.

### Quality control

The first 50,000 individuals included in UKB were genotyped using the Applied Biosystems UK BiLEVE Axiom Array. The remaining were genotyped using the Applied Biosystems UK Biobank Axiom Array. The two array types are equal, and the differences are not of significance. The arrays interrogated 850,000 SNPs in total. To account for potential biases, patients with outlying heterozygosity rates, cryptic relatedness (PIHAT cut-off 0.2) and sex discrepancies in data were excluded. To ensure that only participants with high-quality genomic information were included for analysis, everyone with a genotyping rate of 98% or less were excluded. To ensure that only high-quality genetic variants were left for analyses, a missingness rate of 2% were used as a cut-off point. Lastly, a Minor Allele Frequency (MAF) of > 5% was used, and variants found not to be in Hardy-Weinberg equilibrium were excluded (threshold: 1 × 10^−6^ for both cases and controls).

### Linear Polygenic risk score (PGS) modelling approach

The initial GWAS-analyses were analyzed using a mixed linear model (MLM) approach. GCTA version 1.93 beta for Windows was used to conduct the analyses. The MLM-model was created using fastGWA with a sparse genetic relationship matrix (GRM) with non-imputed data from the UKB. For all the phenotypes analyzed in the respective GWAS, the 100 most significant SNPs were included in the genetic and mixed models. The choice to utilize only the top 100 SNPs was made to optimize the balance between predictive power and keeping the model computational pragmatic. SNPs are referenced using the dbSNP (rs) reference number. The cohorts were split into training/validation and test sets. Relevant GWAS plots, including Manhattan and Quantile-Quantile (QQ) plots were generated using qqman (R version 4.0.2).(13) Performance plots including ROC-AUCs and heatmaps were created using Scikit-learn 1.2.1 (Python 3).(14)

A linear PGS was generated using the logistic regression module as implemented in scikit-learn 1.2.1 for Python 3. Models were created with both L1 (lasso) and L2 (ridge) regularization. Feature importance was determined by coefficients of the SNPs.

### Deep neural network (DNN) modelling approach

All DNN models were implemented using EIR (version 0.1.25-alpha).(15) EIR is a framework that incorporates genetic, clinical, image, sequencing, and binary data for supervised training of deep learning models. A held-out test set was used for all models to obtain a final performance after training and validation. The Cross Entropy loss was employed during training for the classification tasks. All models were trained with a batch size of 64. During training, plateau learning rate scheduling was used to reduce the learning rate by a factor of 0.2 if the validation performance had not improved for 10 steps, with a validation interval of 500 steps. Early stopping was used to terminate training when performance had not improved with a patience of 16 steps. The early stopping criterion was activated after a buffer of 2,000 iterations. All models were trained with the Adam optimizer with a weight decay of 1×10^−4^ and a base learning rate of 1×10^−3^.(16) For the neural network models, we augmented the genotype input by randomly setting 40% of the SNPs as missing in the one-hot encoded array. All DNN models utilize the genome-localnet (GLN) architecture for the genotype feature extraction.(15) The same cohort splits were used as in the linear PGS-approach. Importance of features were determined using SHapley Additive exPlanations (SHAP) values.(17)

## RESULTS

### Cohort

We identified 488,377 patients in the UKB with available genetic and relevant phenotypic data, with 446,180 patients available for analyses after genetic quality measures were applied and were used for both the linear and deep learning modelling approaches.

For the outcomes of interest, 19,704 had a diagnosis of AF, 9,101 had a diagnosis of VTE and 13,757 had a diagnosis of pneumonia overall in the UKB.

### Linear models

#### Atrial fibrillation

Baseline characteristics are listed in table 1. The SNP model reached a ROC-AUC of 60.9% [95% CI, 60.6%-61.0%]. All individuals were classified as not having AF. The clinical model reached a ROC-AUC of 78.7% [95% CI, 78.7%-78.7%] with a recall of 9% and a precision of 53%. The combined model reached a ROC-AUC of 80.1% [95% CI, 80.0%-80.1%] with a recall of 9% and a precision of 57%. All performances are depicted in figure 3A. The SNPs and the associated genes with the highest feature importance are listed in table 4B.

**Table 1:** Baseline characteristics for atrial fibrillation cohort.

| Characteristic | AF<br>N = 19,900 | No AF<br>N = 426,280 |
| --- | --- | --- |
| Age, mean, SD | 62.2 ± 5.9 | 56.4 ± 8.0 |
| Female, N, % | 6,623 (33.2) | 233,351 (54.7) |
| BMI, mean, SD | 27.4 ± 5.4 | 29.1 ± 1 |
| Previous or current smoker,<br>N, % | 9,633 (48.4) | 159,702 (37.5) |
| Previous or current cancer,<br>N, % | 2,307 (11.6) | 35,338 (8.3) |
| Heart failure, N, % | 2,922 (14.7) | 3,252 (0.7) |
| Hypertension, N, % | 12,385 (62.2) | 86,999 (20.4) |

#### Venous thromboembolism

Baseline characteristics for VTE are listed in table 2. The SNP model reached a ROC-AUC of 59.6% [95% CI, 59.0%-59.7%]. All individuals were classified as not having VTE. The clinical model reached a ROC-AUC of 63.4% [95% CI, 63.2%-63.4%]. All individuals were classified as not having VTE. The combined model reached a ROC-AUC of 66.1% [95% CI, 65.7%-66.1%]. All individuals were classified as not having VTE. All performances are depicted in figure 3B. The SNPs and the associated genes with the highest feature importance are listed in table 4D.

**Table 2:** Baseline characteristics for venous thromboembolism cohort.

| Characteristic | VTE<br>N = 9,193 | No VTE<br>N = 436,987 |
| --- | --- | --- |
| Age, mean, SD | 59.7 ± 7.1 | 56.6 ± 8.0 |
| Female, N, % | 4,156 (45.2) | 235,818 (54.0) |
| BMI, mean, SD | 29.4 ± 5.5 | 27.4 ± 4.8 |
| Previous or current smoker,<br>N, % | 3,842 (41.8) | 165,493 (37.9) |
| Previous or current cancer,<br>N, % | 1,403 (15.3) | 36,242 (8.3) |
| Heart failure, N, % | 482 (5.2) | 5,692 (1.3) |
| Hypertension, N, % | 3,906 (42.5) | 95,478 (21.8) |

#### Pneumonia

Baseline characteristics are listed in table 3. The SNP model reached a ROC-AUC of 57.3% [95% CI, 56.5%-57.4%]. All individuals were classified as not having pneumonia. The clinical model reached a ROC-AUC of 69.2% [95% CI, 69.1%-69.2%]. All individuals were classified as not having pneumonia. The combined model reached a ROC-AUC of 70.5% [95% CI, 70.2%-70.6%] with a recall of 0.01% and a precision of 0.4%. The SNPs and the associated genes with the highest feature importance are listed in table 4F. All performances are depicted in figure 3C.

**Table 3:**
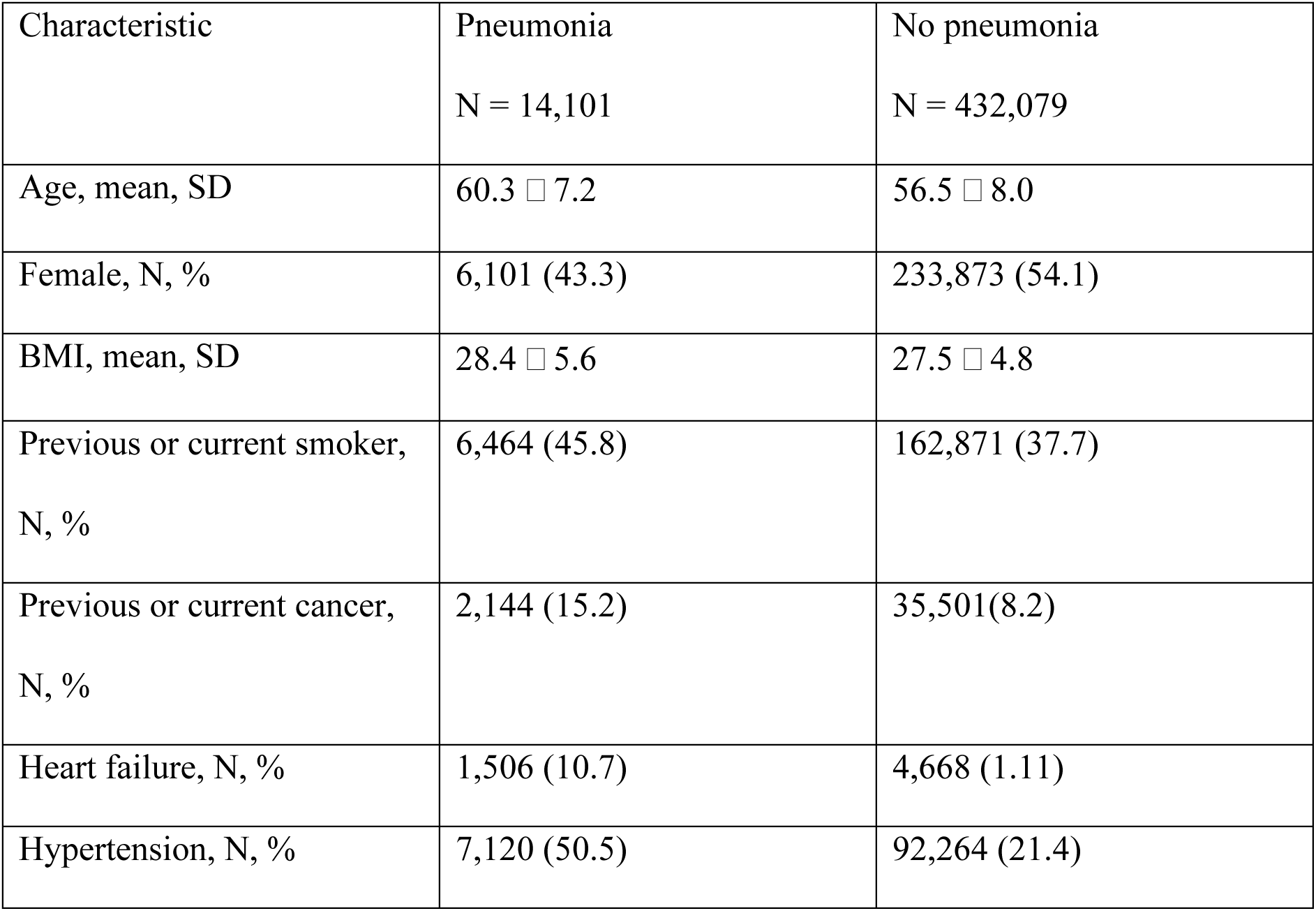
Baseline characteristics for pneumonia.

| Characteristic | Pneumonia<br>N = 14,101 | No pneumonia<br>N = 432,079 |
| --- | --- | --- |
| Age, mean, SD | 60.3 ± 7.2 | 56.5 ± 8.0 |
| Female, N, % | 6,101 (43.3) | 233,873 (54.1) |
| BMI, mean, SD | 28.4 ± 5.6 | 27.5 ± 4.8 |
| Previous or current smoker,<br>N, % | 6,464 (45.8) | 162,871 (37.7) |
| Previous or current cancer,<br>N, % | 2,144 (15.2) | 35,501 (8.2) |
| Heart failure, N, % | 1,506 (10.7) | 4,668 (1.11) |
| Hypertension, N, % | 7,120 (50.5) | 92,264 (21.4) |

### Deep learning models

#### Atrial fibrillation

The SNP model reached a ROC-AUC of 59.9% [95% CI, 58.6%-61.3%] in the test set. Recall was 36.9% and precision was 9.3%. The clinical model reached a ROC-AUC of 78.8% [95% CI, 77.8%-79.8%] with recall and precision of 72.0% and 13.5%, respectively. The combined model reached a ROC-AUC of 79.4% [95% CI, 78.8%-80.5%] with a recall and precision of 74.8% and 13.5%, respectively. The SNPs and the associated genes with the highest feature importance are listed in table 4A. All performances are depicted in figure 3A. Hard predictions are depicted in figure 1A. ROC-AUC development is depicted in figure 2A.

**Figure 1A:**
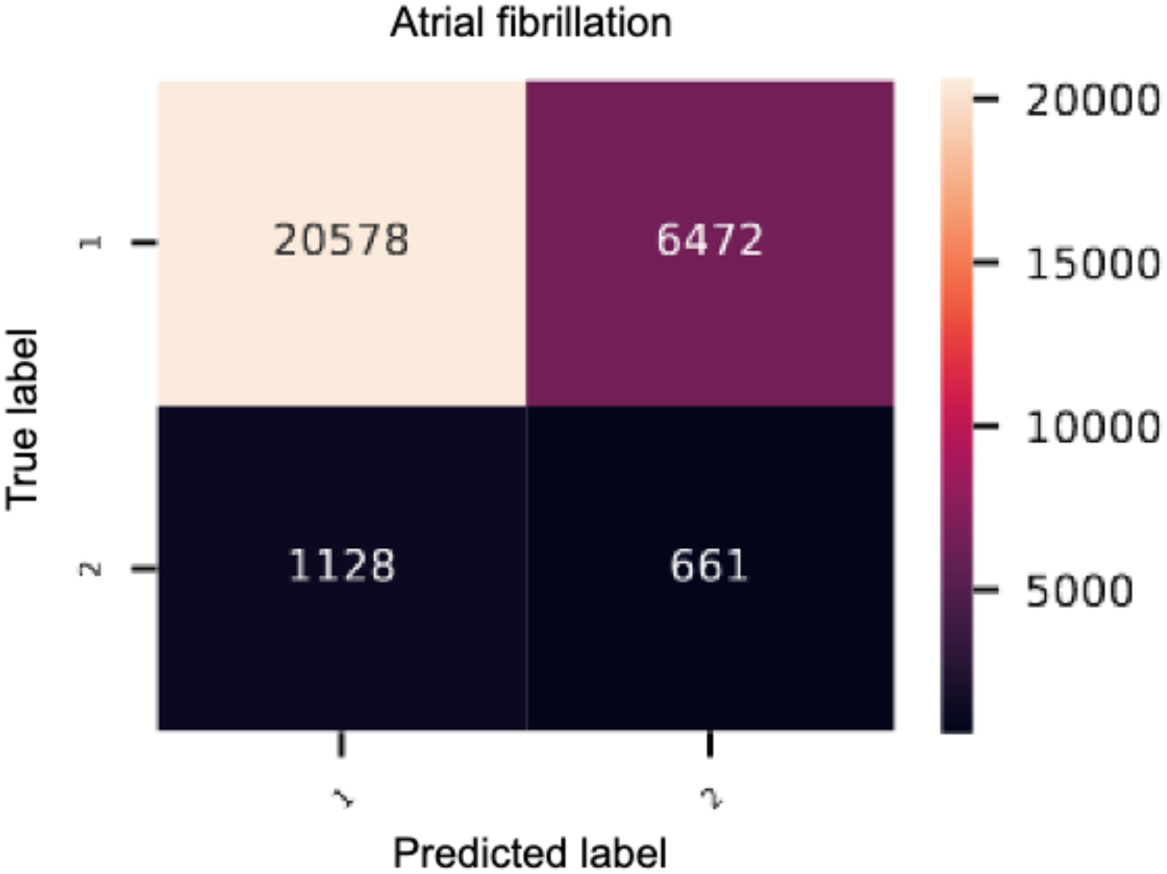
Heatmap of the genetic deep learning-model of atrial fibrillation. 1 = controls, 2 = cases

**Figure 2A:**
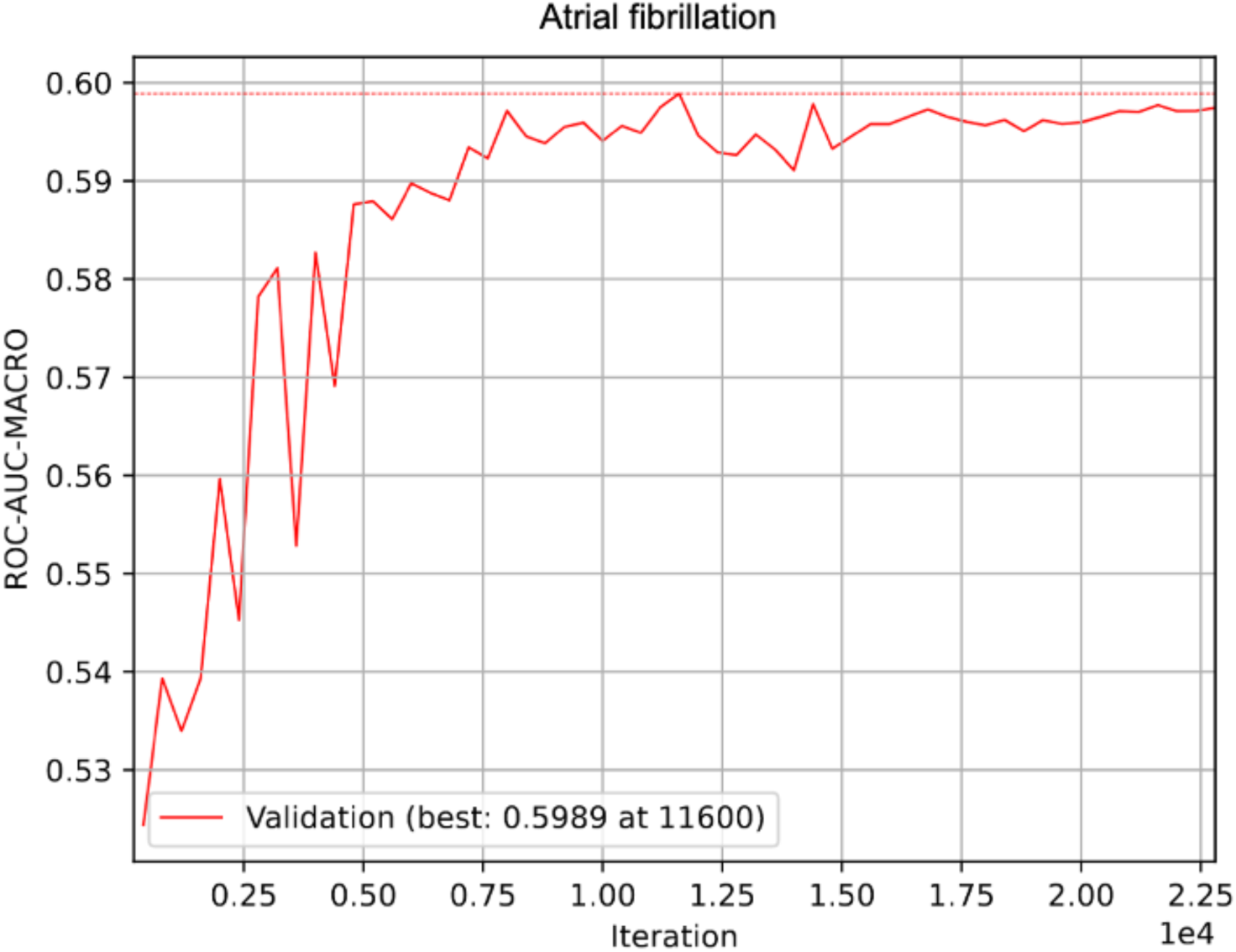
ROC-AUC development of the genetic GLN-model of atrial fibrillation

**Figure 3A:**
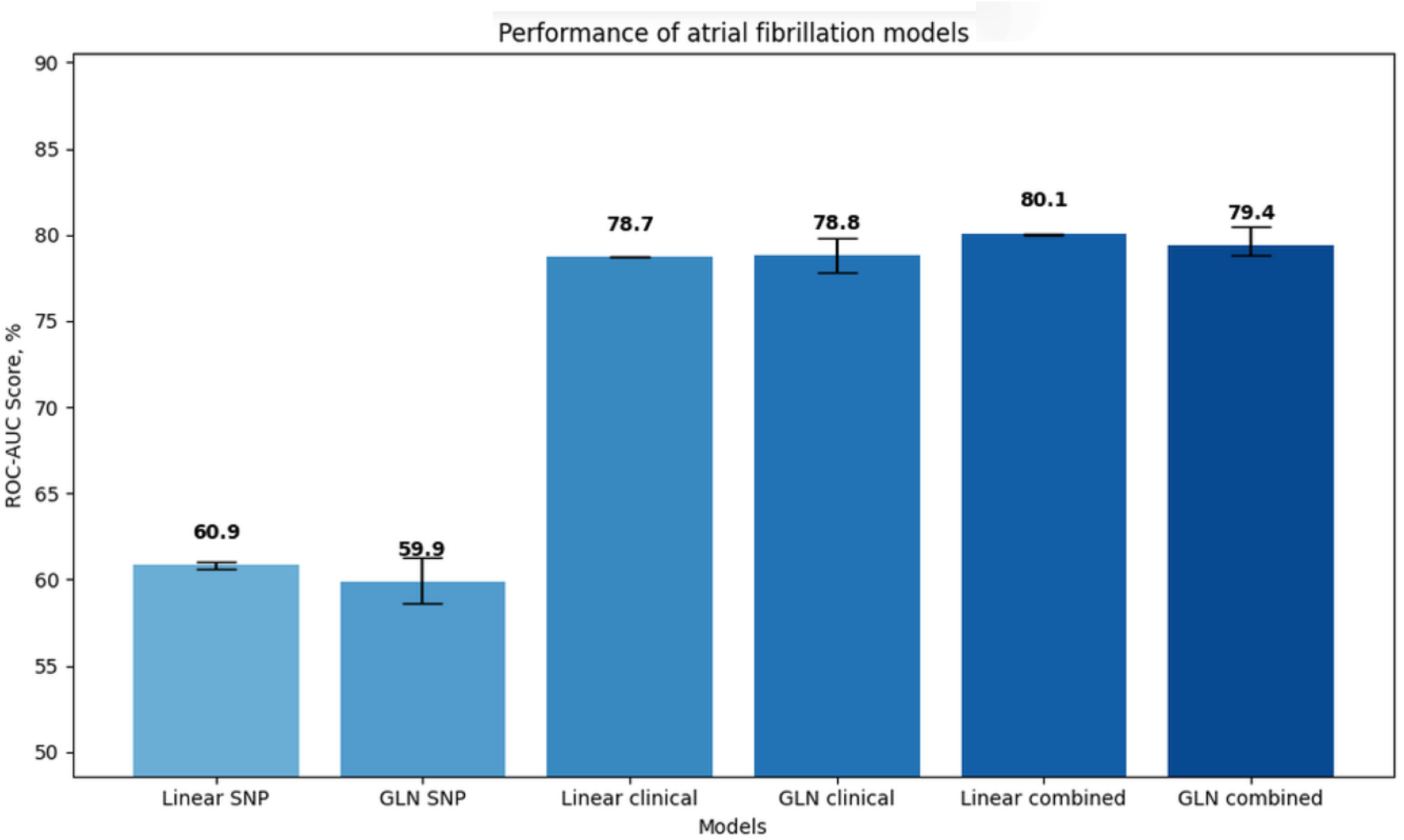
Bar plot of ROC-AUCs of all atrial fibrillation models

**Table 4A:**
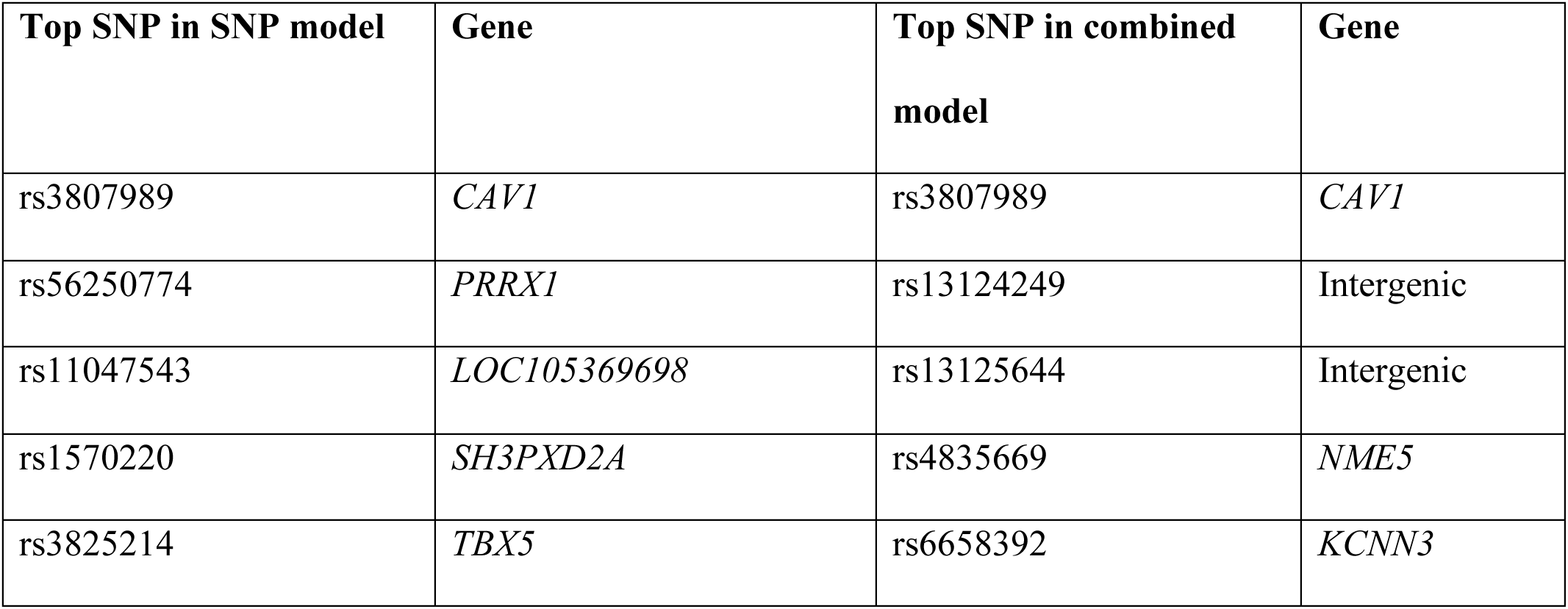
Table of the SNPs with the highest feature importance for the single nucleotide polymorphism (SNP), and combined atrial fibrillation GLN-models.

**Table 4B:**
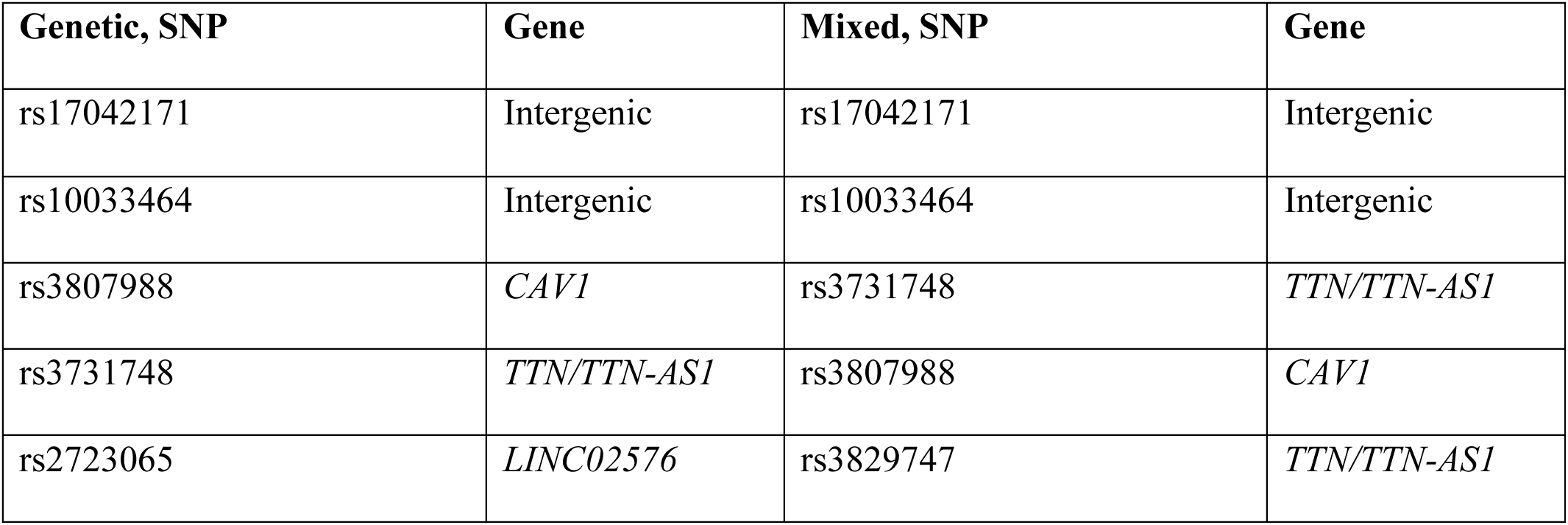
Table of the SNPs with the highest feature importance for the genetic, and mixed atrial fibrillation linear models.

**Table 4C:** Table of the SNPs with the highest feature importance for the single nucleotide polymorphism (SNP), and combined venous thromboembolism GLN-models.

| Top SNP in SNP model | Gene | Top SNP in combined model | Gene |
| --- | --- | --- | --- |
| rs505922 | *ABO* | Rs6993770 | *ZFPM2* |
| rs657152 | *ABO* | Rs687621 | *ABO* |
| rs8176740 | *ABO* | Rs75112989 | *ATP1B1* |
| rs7868232 | Intergenic | Rs3746438 | *MYH7B* |
| rs581107 | *ABO* | Rs56103207 | *TSPAN15* |

**Table 4D:** Table of the SNPs with the highest feature importance for the genetic, and mixed Venous thromboembolism linear models.

| Genetic, SNP | Gene | Mixed, SNP | Gene |
| --- | --- | --- | --- |
| rs4524 | *F5* | rs4524 | *F5* |
| rs6030 | *F5* | rs6120849 | *EDEM2* |
| rs6120849 | *EDEM2* | rs6030 | *F5* |
| rs75112989 | *ATP1B1* | rs75112989 | *ATP1B1* |
| rs6050 | *FGA* | rs507666 | *ABO* |

**Table 4E:**
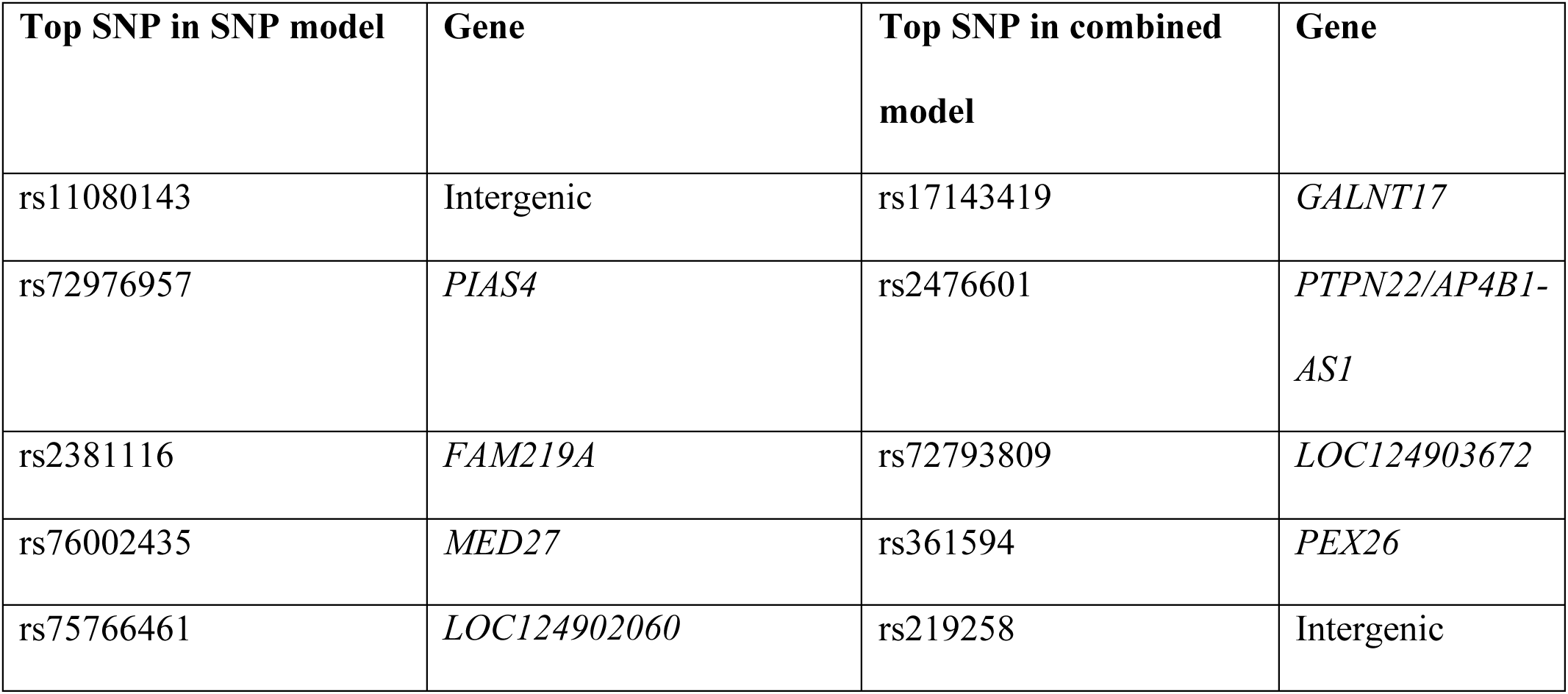
Table of the SNPs with the highest feature importance for the single nucleotide polymorphism (SNP), and combined pneumonia GLN-models.

**Table 4F:**
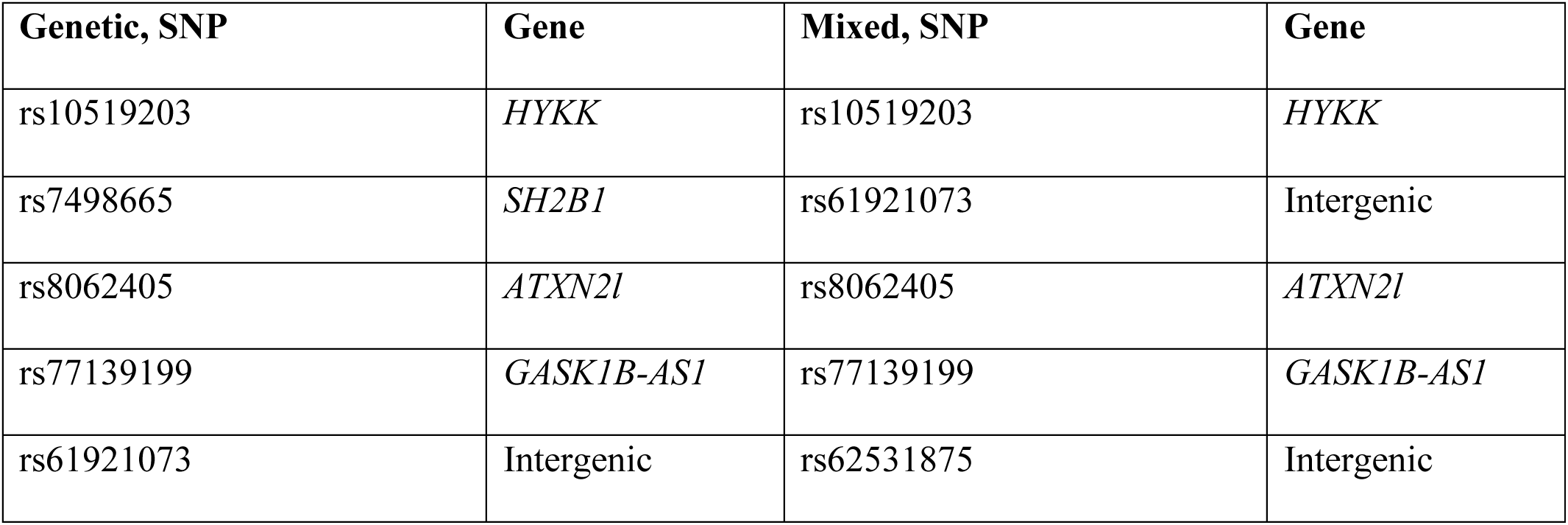
Table of the SNPs with the highest feature importance for the genetic, and mixed pneumonia linear models.

#### VTE

The SNP model reached a ROC-AUC of 60.0% [95% CI, 57.8%-61.8%] with a recall of 50.8% and precision of 4%. The clinical model reached a ROC-AUC of 63.2% [95% CI, 61.2%-65.0%] with a recall and precision of 67.5% and 4.0%, respectively. The combined model reached a ROC-AUC of 65.4% [95% CI, 63.6%-67.2%] with a recall and precision 68.8% and 4.0%, respectively. The SNPs and the associated genes with the highest feature importance are listed in table 4C. All performances are depicted in figure 3B. Hard predictions are depicted in figure 1B. ROC-AUC development is depicted in figure 2B.

**Figure 1B:**
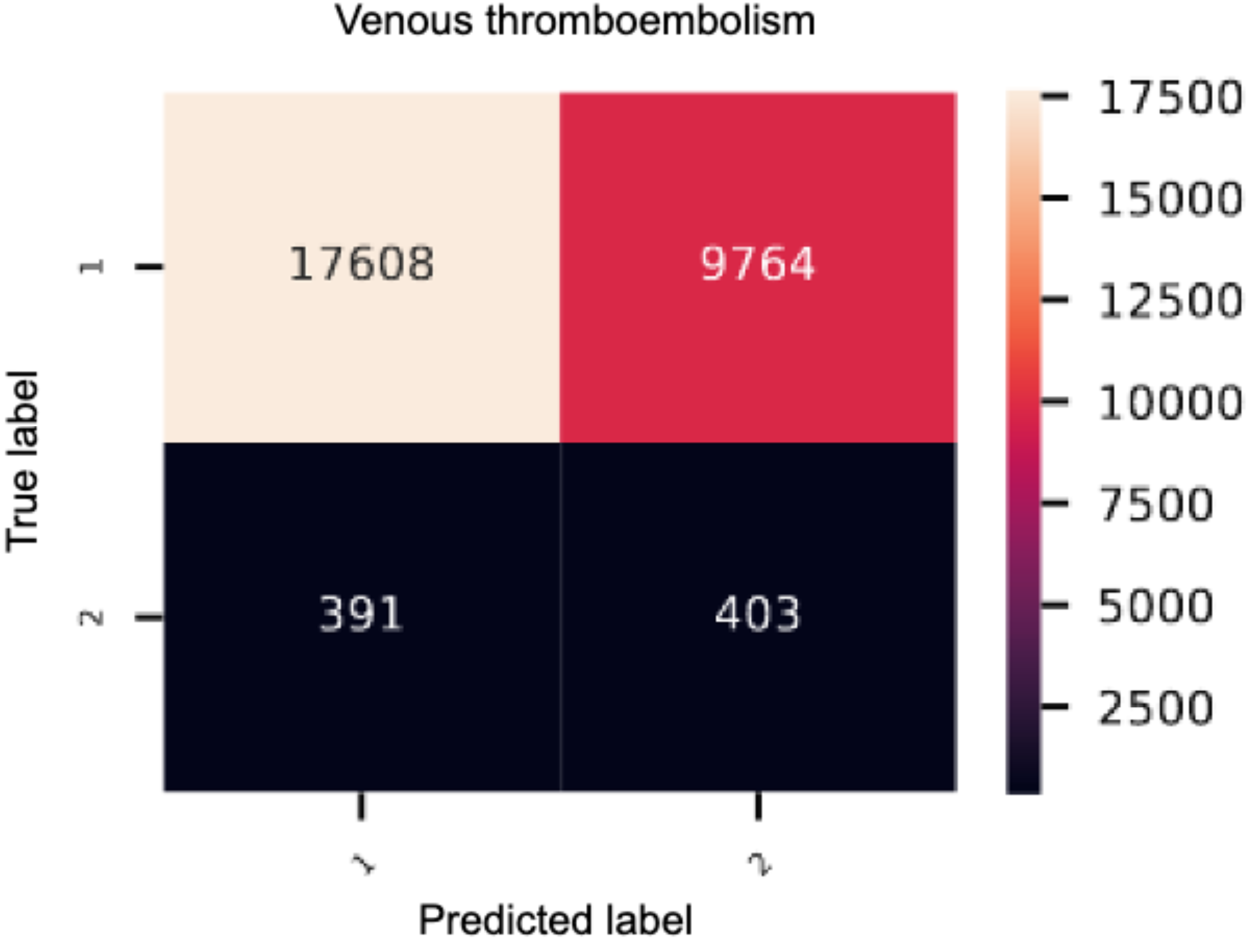
Heatmap of the genetic deep learning-model of venous thromboembolism. 1 = control, 2 = cases. VTE: venous thromboembolism

**Figure 2B:**
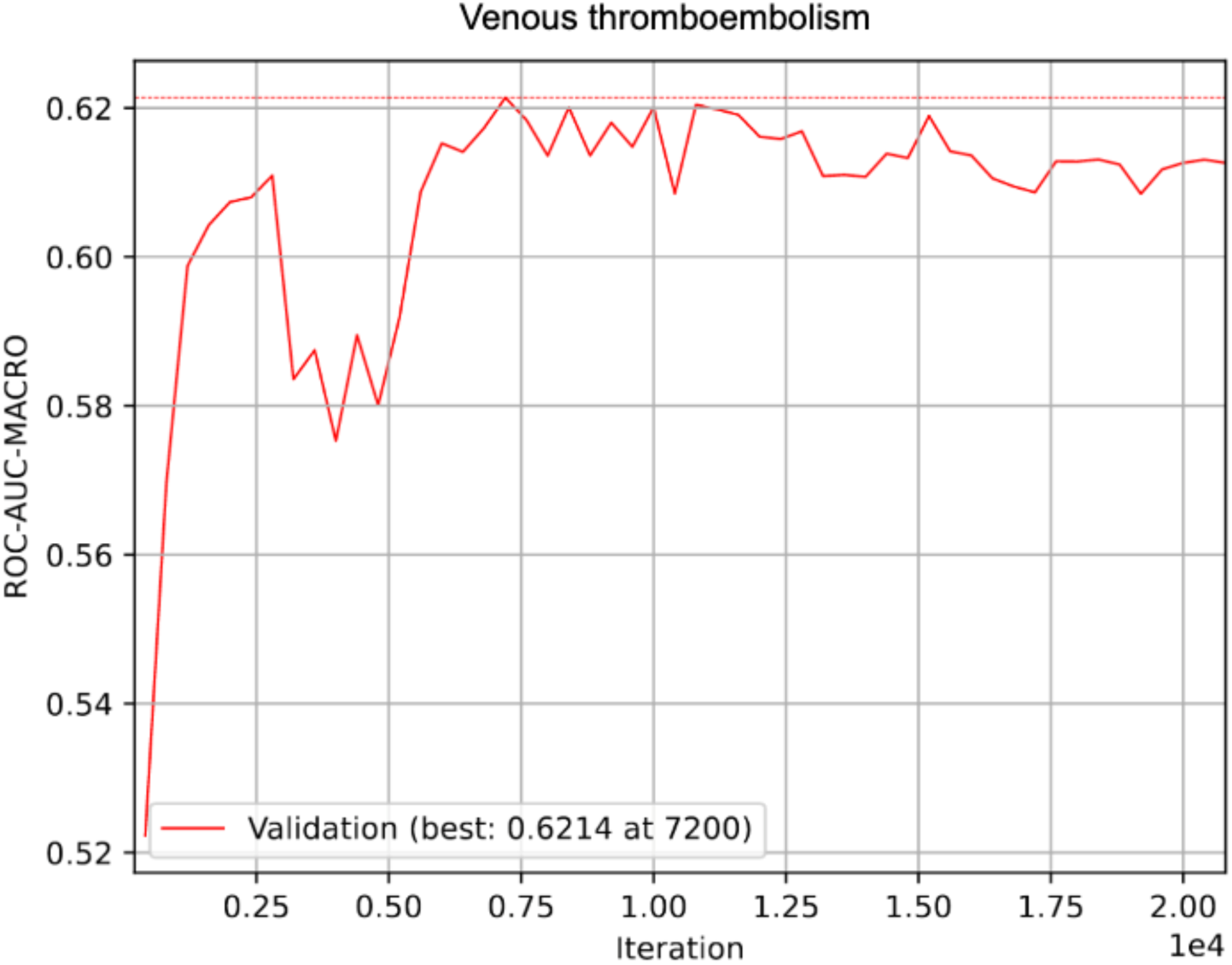
ROC-AUC development of the genetic GLN-model of venous thromboembolism

**Figure 3B:**
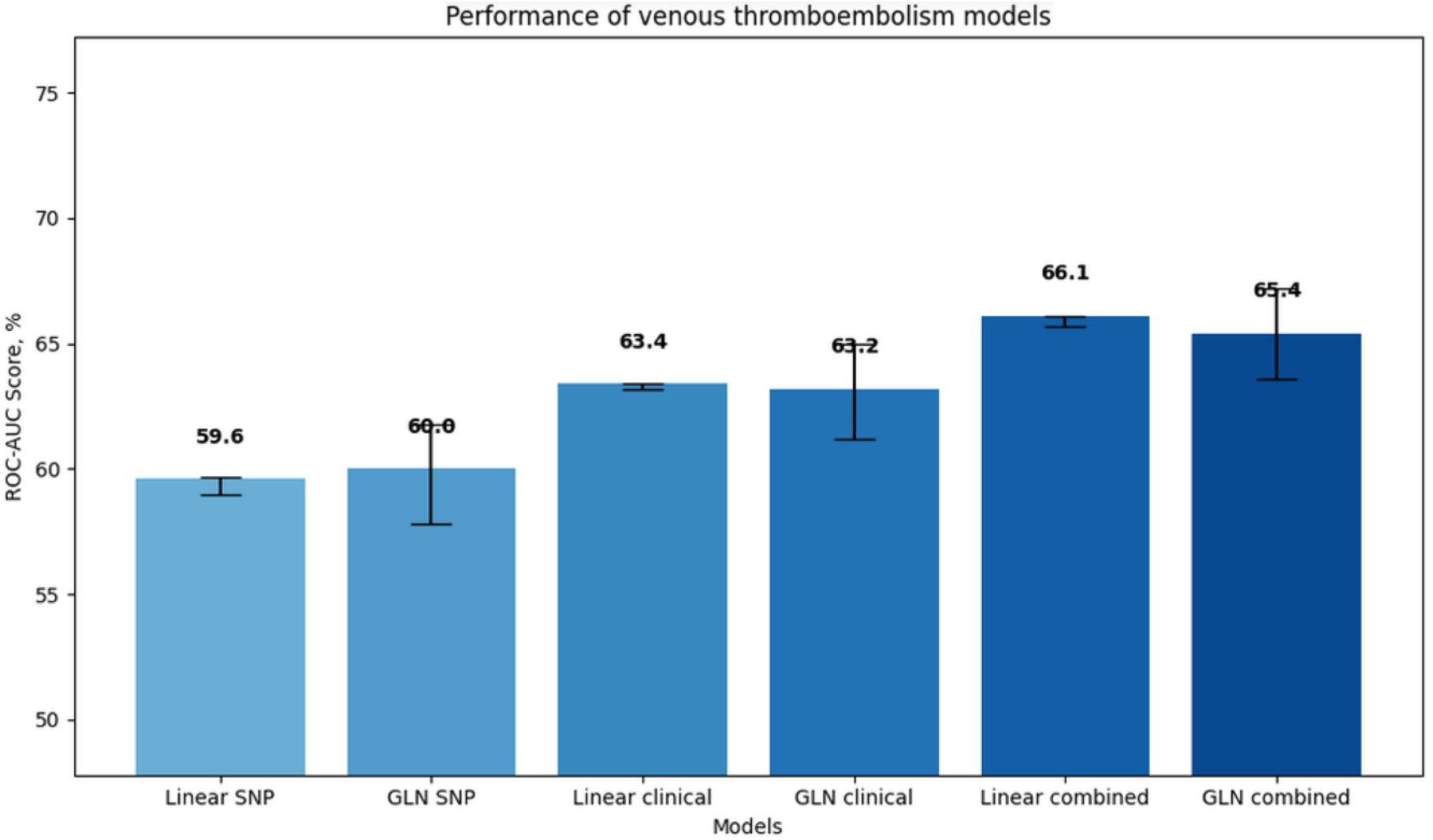
Bar plot of ROC-AUCs of all venous thromboembolism models

#### Pneumonia

The SNP model reached a ROC-AUC of 55.5% [95% CI, 54.1%-56.9%] with a recall of 55.0% and precision of 5%. The clinical model reached a ROC-AUC of 69.7% [95% CI, 68.5%-70.8%] with a recall and precision of 67.7% and 7.4%, respectively. The combined model reached a ROC-AUC of 69.9% [95% CI, 68.7%-71.0%] with a recall and precision of 70.1% and 7.3%, respectively. The SNPs and the associated genes with the highest feature importance are listed in table 4E. All performances are depicted in figure 3C. Hard predictions are depicted in figure 1C. ROC-AUC development is depicted in figure 2C.

**Figure 1C:**
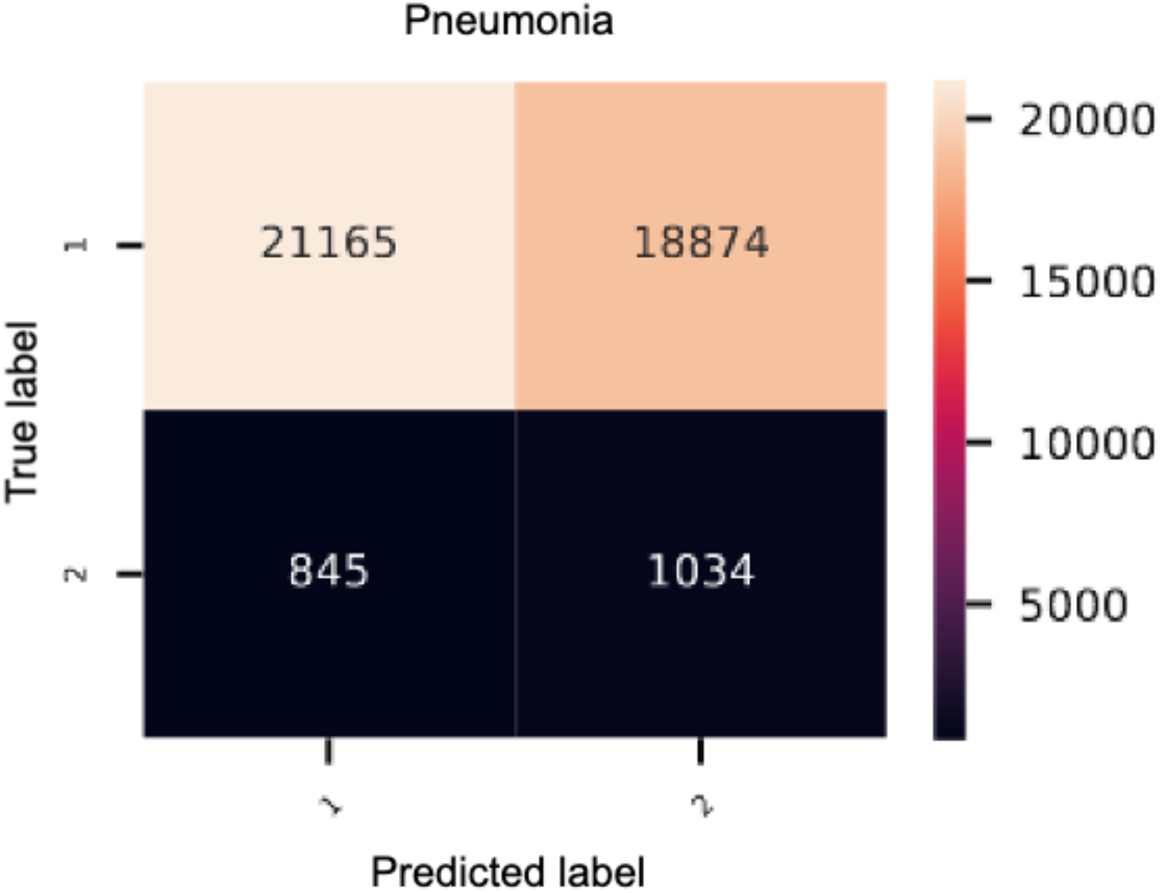
Heatmap of the genetic deep learning model of bacterial pneumonia. 1 = control, 2 = cases.

**Figure 2C:**
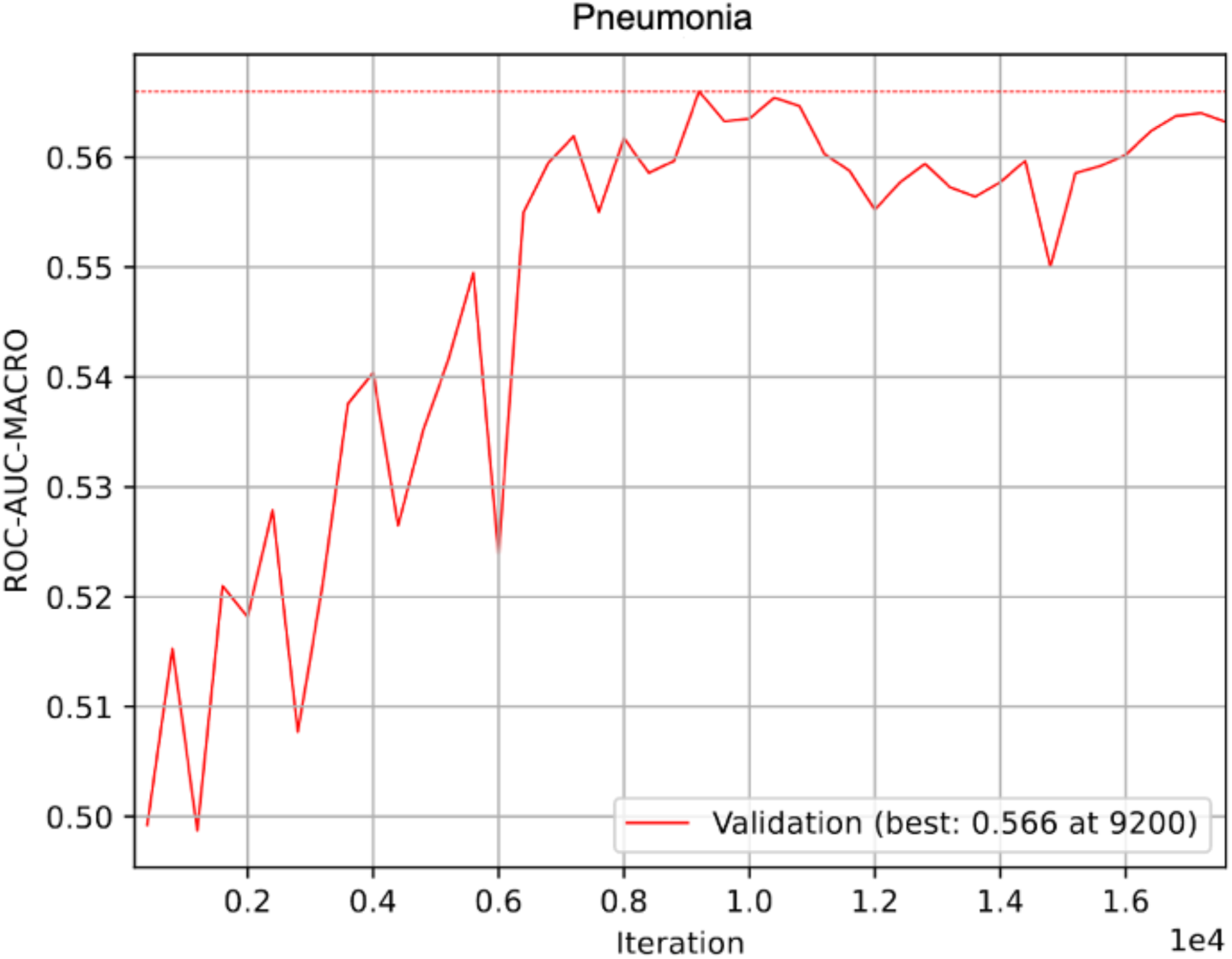
ROC-AUC development of the genetic GLN-model of bacterial pneumonia

**Figure 3C:**
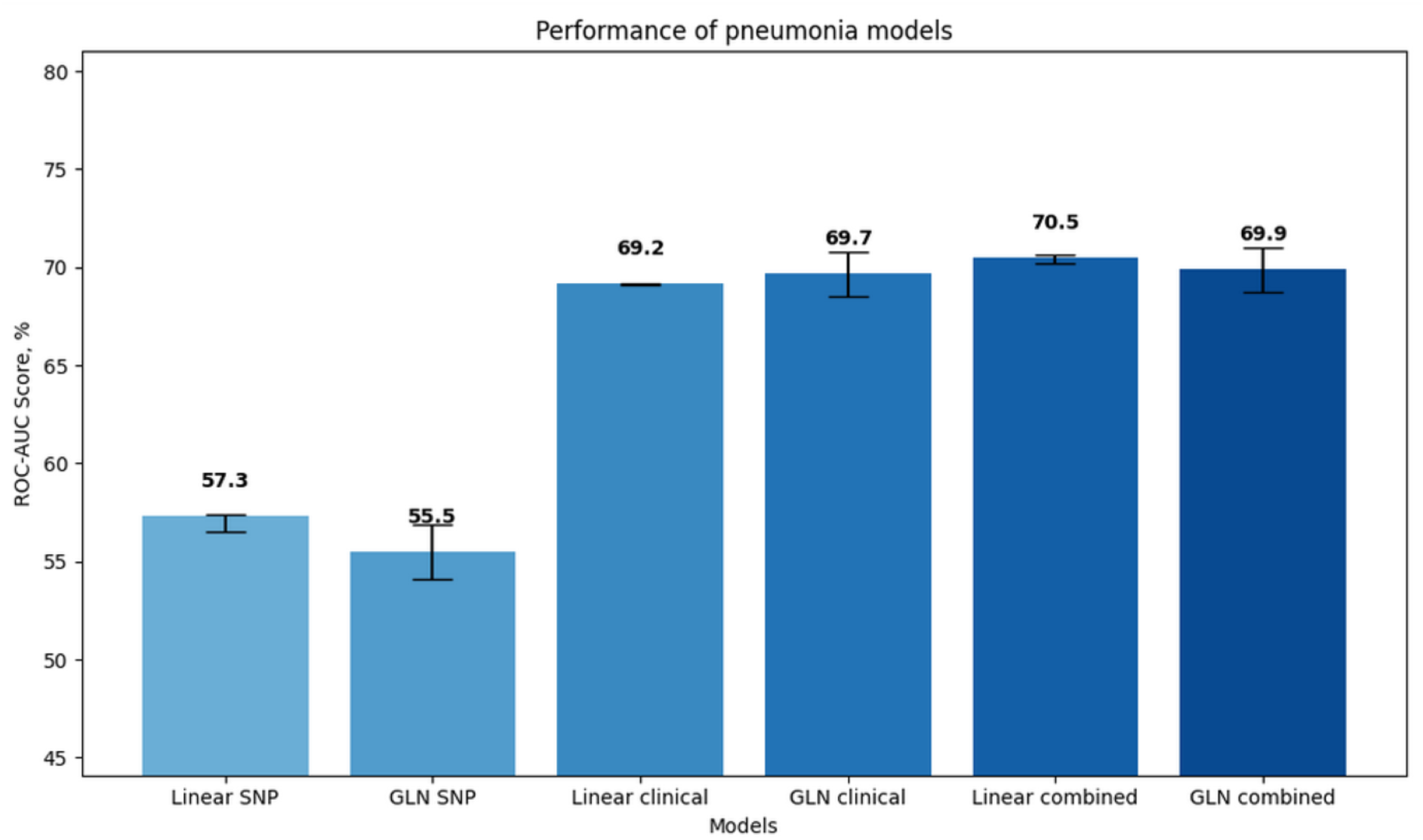
Bar plot of ROC-AUCs of all pneumonia models

**Figure 4:**
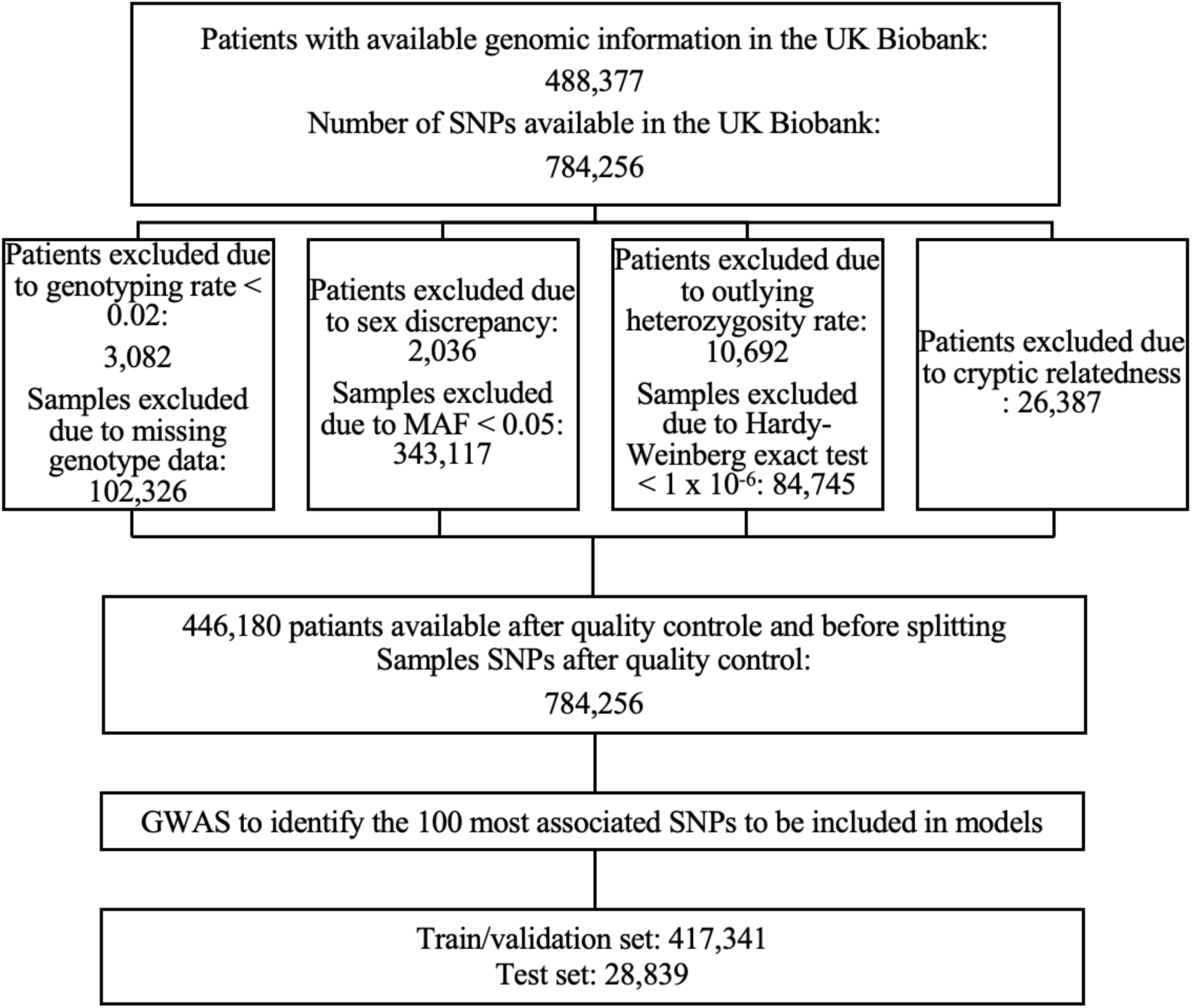
Selection and quality control steps of individuals and SNPs in the UKB

## DISCUSSION

In this study, we assessed the performance of linear and deep learning models including genotypic information on specific phenotypes relevant for pAFLI, pVTE and postoperative pneumonia. Overall, we found that adding SNP data to clinical risk prediction models enhanced the predictive power, and that the GLN approach seemed superior to a legacy linear risk prediction approach.

### Modelling approaches

All three SNP linear models failed to make any meaningful hard predictions, as they classified all individuals in the cohorts as not having the disease in question. However, the separations were roughly similar to the GLN-models, as demonstrated by similarity in ROC-AUC performance, and the lack of positive predictions may be due to imbalanced data and skewed threshold for hard predictions. The GLN-models performed better on recall and precision, and it was able to classify positives correctly with just genomic information. Given that the linear and GLN models utilize distinct tuning parameters for hard predictions, a direct comparison of recall and precision may not be critically significant. However, as hard predictions are necessary in a clinical setting, a discussion is still warranted. Precision-recall curves are depicted in supplementary materials.

It is exemplified for AF, where the genetic linear model had a ROC-AUC of 60.9% [95% CI 69.6%-61.0%] while the GLN had a ROC-AUC of 59.9% [95% CI 5.8.6%-61.3%]. The recall, however, was 0% and 36.9%, respectively. As the linear model performed very poorly in terms of recall, the result is likely to be the result of the imbalanced data and failure to capture feature relevance and possibly non-linearity and would need optimization before any form of utilization for positive prediction in a clinical setting. On the contrary, the GLN-model had a recall of 36.9% and therefore identifies around one third of cases correctly, which heightens the likelihood of clinical meaningful utilization considering only SNPs were included in the model. However, the precision was calculated low at 4%, which would lead to overdiagnosis and possibly overtreatment. If used in clinical practice, it is therefore of paramount importance that any possible intervention would carry little to no risk of harm. It, however, cannot be ruled out that the differences between the models are not due to an inherent predictive advantage in the GLN-model, but simply due to different hyperparameter tuning.

When combining SNP and clinical data for all phenotypes in question, we observed a trend towards better performance compared with SNP or clinical data only models, although most confidence intervals were overlapping with clinical model performances. This thus indicates that limited performance gains could obtained by combining genetic and clinical data and may suggest that genotype effects may already in part be captured by diagnoses codes.

Alternatively, the lack of performance improvement could be affected by limited study power due to factors such as lack of correct PC diagnoses codes, a problem often encountered when administrative codes are used for PC curation.(18)

GLN based models did, however, outperform linear approaches in terms of recall performance, might indicate that the ability to capture the effects of non-linear genetic traits on the overall phenotype, may be possible through this modelling approach.

### Identified Single Nucleotide Polymorphisms

In the GLN-model, rs3807989 was the most activated SNP in regards of classifying individuals with AF. It is an intron variant in *CAV1* which codes for a main component in caveolae plasma membrane and further acts as a tumor suppressor.(19, 20) It has been associated with a large variety of diseases including AF in numerous populations.(21, 22) Interestingly, the prevalence of the reference and risk allele is roughly equal which suggests the possibility of a relatively new mutation or that the risk variant has a different functional advantage which balances the selection. The most highly activated SNP in the GLN-model with importance for classifying patients for not having AF was rs17042081, a variant near 4q25, which has been extensively associated with AF in a variety of populations and in close proximity to *PITX2*.(23, 24), The variants most highly associated with AF in the linear model was rs17042171, also a variant near 4q25. The alternative allele has worldwide prevalence of up to 16% and 13% in the European population, which makes the risk variant very common, although not equal to the reference allele suggesting a negative selection pressure of the risk allele.(25) As the testing set consists of purely surgical patients, it is not unexpected that variants near 4q25 are important for the models, as the same region was the only one associated with postoperative AF in a recent GWAS-analysis from our group.(7)

The difference in which variants show importance for the GLN and linear model, respectively, and that the GLN-models in general performed significantly better in recall compared with the matching linear, shows that non-linear interactions between genes which are potentially of great importance in the risk of a particular trait. Other explanations include non-linear effects in non-genetic features, such as age and sex, or dominant/recessive effects of the SNPs in question.

When exploring pathways and interactions for the most highly activated genes in online repositories such as the Reactome Pathway Database and BioGRID, it appears that none of the genes have previously been described to be in a direct pathway or in any kind of interaction. Interestingly, Gao et al. showed that the level of caveolin-1 determines the level of product of *KCNN1* which previously has been highly associated with AF in several GWAS-studies.(21, 26–28)

The SNPs with highest importance for classifying VTE in the GLN-model was rs505922, an intron variant in *ABO*.(29) The variant has previously been associated with VTE.(6) The ABO blood group antigen genes are amongst the most heavily associated with VTE, and it has a biological plausible explanation, thus it is expected that specific variants within these genes would play a significant role in predictive models for VTE risk.

The SNP that had the highest feature importance in the GLN-model for classifying bacterial pneumonia was rs11080143. However, as the model performed poorly in terms of the overall accuracy, there is a high likelihood that the highest activated variants are not due to genuine, biological phenomena and interactions, but rather due to chance alone. This sentiment is further supported by the fact that rs11080143 has no reported clinical significance in the literature and does not lie close to any biological meaningful genes. One downstream gene, *KSR1*, has been associated with different malignancies including breast adenocarcinoma and thyroid cancer.(30, 31) To our knowledge, *KSR1* has not been associated directly with lung cancer, but considering the protein product has a positive downstream signaling function of the RAS/MAPK pathway, and the association with other cancers, a connection seems likely. Although a history of cancer was included as a covariate in the initial GWAS and in both models, it cannot be ruled out that the phenotype and models are confounded by occult lung malignancy.

In the linear model, rs10519203 was the most associated with classifying pneumonia. It is an intron variant in *HYKK* and has previously been associated with lung cancer and smoking behavior, which may indicate the basis for its ability to classify pneumonia, and not an inherent increased risk to infection.(32, 33)

We again observed a discrepancy between the variants with highest feature importance between the models, which suggests that complex non-linear effects may exist between the genes in question activated by the GLN-model. It should be noted that while all the phenotypes of interest in this study are complex diseases, we find it likely that the susceptibility to bacterial pneumonia is less driven by genetics compared with AF and VTE.

Although certain genetic variants have been associated pneumonia susceptibility, the genetic landscape has not been explored to the same extent as with AF and especially VTE. Consequently, the ratio of importance of the input variants compared with the clinical factors included is likely lower compared with AF and VTE, and we anticipate that future research will highlight the importance of genetics compared with clinical factors in predictive modelling.

### Potentials for clinical use

As these analyses were specifically made on phenotypes relevant for the postoperative course of surgical patients, it is of utmost importance that the models in question can be validated and potentially optimized in a specific surgical cohort. Above all, this will ascertain the utilization on this specific population, and it will further establish a foundation for the investigation into if the models are able to improve the outcomes for the phenotypes in question or be of prophylactic benefit. Further, as multiple surgical risk predictors built on clinical data already exist, investigate whether the addition of genetic data will enhance the predictability of such models could offer a promising pathway for increasing the model performances further. It is key to establish methods to improve prediction, as the current standard of models fail to demonstrate any clear clinical benefit compared with standard practice.(34) Although multiple factors account for the current limited applicability, including lacking external validity and variance in the retro- and prospective data, a lack of important factors such as genetics may also be of significance. Further, as we present a model where a deep learning framework specifically made to incorporate genetics with clinical variables that performs better compared with a linear PGS in terms of recall and precision, it is important to consider the quality of the used software as well as the pragmatic applicability of the models in question in a real-life scenario. At this time, neither model have applicability if incorporating only the top 100 SNPs, as determined by the poor accuracy performance.

### Limitations

This study has limitations. First, all phenotypes were established using only ICD-codes which may have a low accuracy for the phenotypes in question. We suspect especially bacterial pneumonia to have an overall low accuracy due to the high hospital incidence and difference in presentation as well differences in the microbiological organism and treatments. This generates a heterogenous group which lowers the predictability and clinical utility. Further, we assumed that the one hundred most significant SNPs from an initial GWAS for the phenotype in question would be of interest, although this was an arbitrary choosing due to the need to find an optimum between predictive power and computational efficacy. Other SNPs may also be of importance, and using a different set or potentially the entire genome has the potential to achieve similar or even better genetic predictability, although the latter would be too computational costly and of less clinical utility. A significant challenge in our study is the imbalanced data, which especially proved problematic in the linear models which all had a recall of 0. Larger cohorts, or a more balanced dataset may improve this.

### Conclusion

In conclusion, we present predictive models on surgery relevant phenotypes incorporating a small sample of genetic variants. Overall, GLN-based models performed equally when compared with linear models based on the AUC metric. However, recall and precision were better in the GLN-based model, making them more useful in a potential clinical setting. Further, different SNPs were important for the same phenotypes between models suggesting importance off non-linear interactions. Lastly, in a comparison between clinical models with and without inclusion of SNPs, the inclusion of genetic data seemed to increase the accuracy, albeit with overlapping confidence intervals. This is a preliminary report assessing the utility of using a small sample of SNPs for clinical risk prediction. Future research needs to validate models in surgical cohorts and assess the utility of incorporating genetics and clinical variables in predictive models to improve surgical outcomes.

## Data Availability

Data is not publicly available but can be applied for at https://www.ukbiobank.ac.uk/enable-your-research/apply-for-access. Analytic methods will be made public at github.com at request. Requests to access these datasets should be directed to https://www.ukbiobank.ac.uk/enable-your-research/apply-for-access.

---

**Supplementary table 1:** List of ICD-9 and ICD-10 codes used for the phenotypes in question.

|  |  |
| --- | --- |
| ICD-9 codes for atrial fibrillation | 4273 |
| ICD-10 codes for atrial fibrillation | I48, I480, I481, I483, I484, I489 |
| ICD-9 codes for venous thromboembolism | 4151, 4511, 4512, 4519, 4531, 4532, 4534, 4538, 4539, 4534, 4531, 4532, 4539 |
| ICD-10 codes for venous thromboembolism | I260, I269, I801, I802, I803, I808, I809, I820, I821, I822, I823, I828, I829, O082, O223, O871, O882, I81 |
| ICD-9 codes for pneumonia | 4810, 4820, 4821, 4823, 4824, 4828, 4829, 4830, 4831, 4838, 4840, 4841, 4843, 4835, 4836, 4847, 4848, 4850, 4860 |
| ICD-10 codes for pneumonia | J13, J14, J150, J151, J152, J153, J154, J155, J156, J157, J158, J159, J16, J160, J168, J170, J172, J173, J178, J180, J181, J182, J188, J189, J851 |

**Supplementary table 2:**
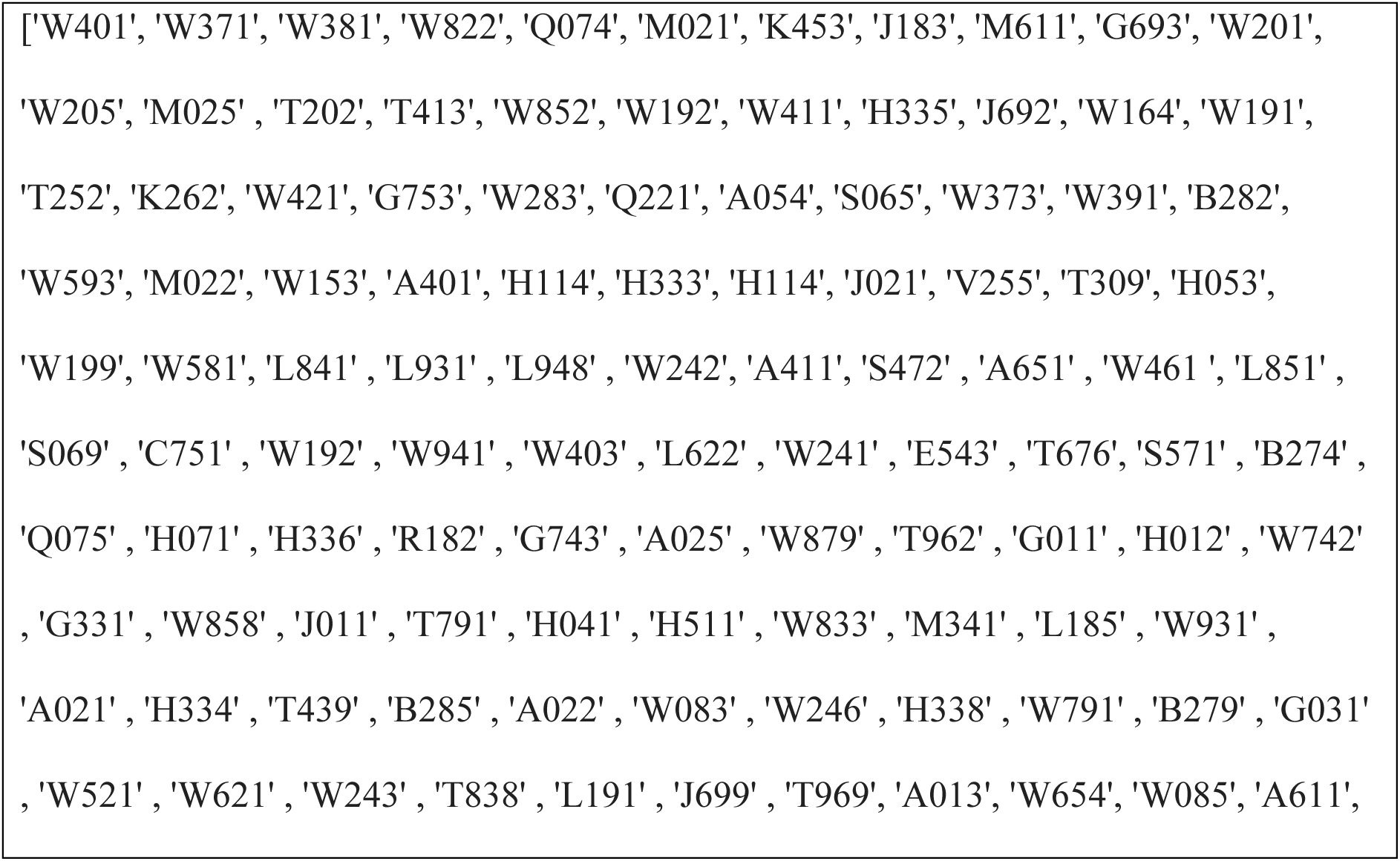
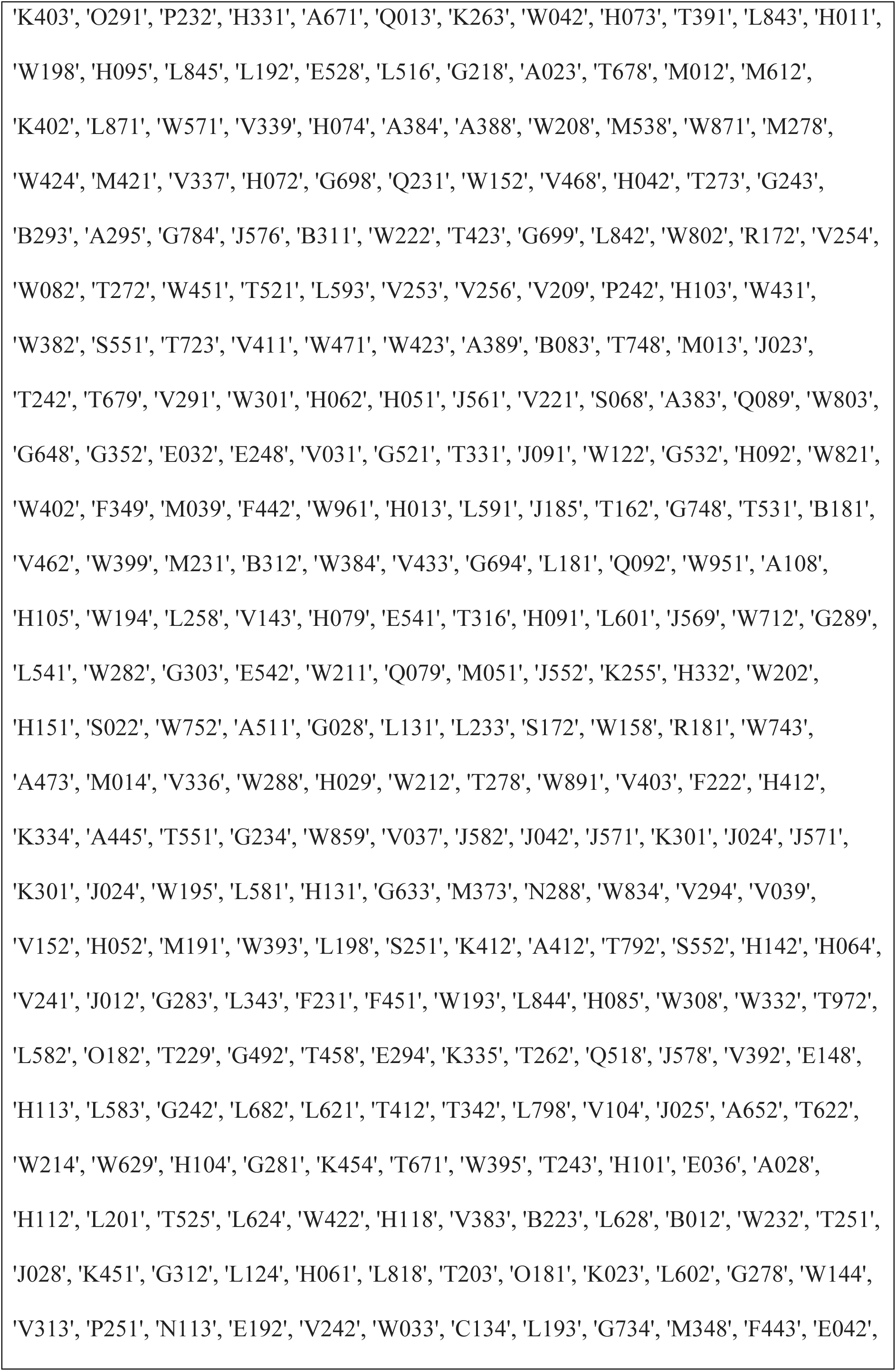
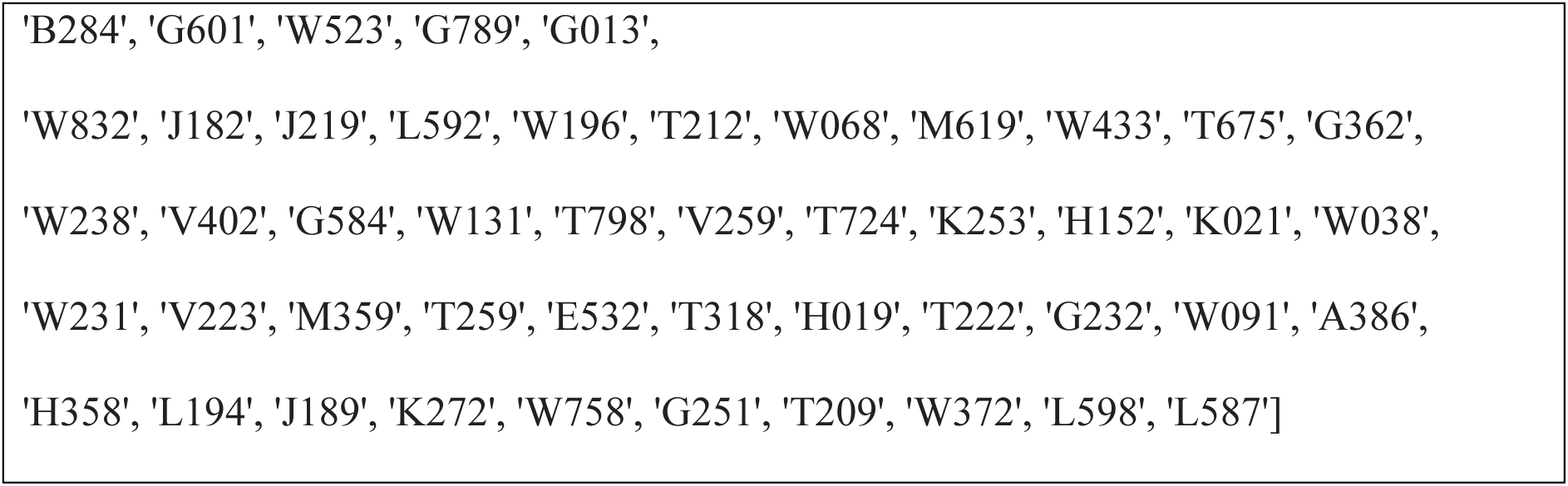
List of OPCS-4 codes used to define surgery.

**Supplementary table 3:** Demographics and comorbidities with ICD-codes.

|  |  |
| --- | --- |
| Demographics | **Age** (UKB code field: 21022-0.0)<br>**Sex** (UKB code field: 31-0.0) |
| Comorbidities | **Previous or current smoker** (UKB code field: 1249-0.0, 1249-1.0, 1249-2.0, 1249-3.0)<br>**Hypertension** (ICD-10: I10, I100, I101, I102, I103, I104, I105, I106, I107, I108, I109, I13, I130, I131, I132, I133, I134, I135, I136, I137, I138, I139, I15, I150, I151, I152, I153, I154, I155, I156, I157, I158, I159)<br>**Cancer** (UKB code field: 2453-0.0, 2453-1.0, 2453-2.0, 2453-3.0 ), heart failure (ICD-10: I50, I501, I502, I5020, I5021, I5022, I5023, I503, I5030, I5031, I5032, I5033, I504, I5041, I5042, I5043, I508, I5081, I50810, I50810, I50811, I50812, |
|  | I50813, I50814, I5082, I5083, I5084, I5089,<br>I509)<br>**BMI** (UKB code field: 21001-0.0, 21001-1.0, 21001-2.0, 21001-3.0. |

## REFERENCES

1. Ozgediz D, Jamison D, Cherian M, McQueen K. The burden of surgical conditions and access to surgical care in low- and middle-income countries. Bull World Health Organ. 2008;86(8):646–7.

2. Weiser TG, Haynes AB, Molina G, Lipsitz SR, Esquivel MM, Uribe-Leitz T, et al. Estimate of the global volume of surgery in 2012: an assessment supporting improved health outcomes. Lancet. 2015;385 Suppl 2:S11.

3. Dencker EE, Bonde A, Troelsen A, Varadarajan KM, Sillesen M. Postoperative complications: an observational study of trends in the United States from 2012 to 2018. BMC Surg. 2021;21(1):393.

4. Bertsimas D, Dunn J, Velmahos GC, Kaafarani HMA. Surgical Risk Is Not Linear: Derivation and Validation of a Novel, User-friendly, and Machine-learning-based Predictive OpTimal Trees in Emergency Surgery Risk (POTTER) Calculator. Ann Surg. 2018;268(4):574–83.

5. Bilimoria KY, Liu Y, Paruch JL, Zhou L, Kmiecik TE, Ko CY, et al. Development and evaluation of the universal ACS NSQIP surgical risk calculator: a decision aid and informed consent tool for patients and surgeons. J Am Coll Surg. 2013;217(5):833–42 e1-3.

6. M AC, Bonde A, Sillesen M. An assessment of the effect of the genotype on postoperative venous thromboembolism risk in 140,831 surgical patients. Ann Med Surg (Lond). 2021;71:102938.

7. Christensen MA, Bonde A, Sillesen M. Genetic risk factors for postoperative atrial fibrillation—a nationwide genome-wide association study (GWAS). Frontiers in Cardiovascular Medicine. 2023;10.

8. Gaudino M, Di Castelnuovo A, Zamparelli R, Andreotti F, Burzotta F, Iacoviello L, et al. Genetic control of postoperative systemic inflammatory reaction and pulmonary and renal complications after coronary artery surgery. J Thorac Cardiovasc Surg. 2003;126(4):1107–12.

9. Kolek MJ, Muehlschlegel JD, Bush WS, Parvez B, Murray KT, Stein CM, et al. Genetic and clinical risk prediction model for postoperative atrial fibrillation. Circ Arrhythm Electrophysiol. 2015;8(1):25–31.

10. Bonde A, Varadarajan KM, Bonde N, Troelsen A, Muratoglu OK, Malchau H, et al. Assessing the utility of deep neural networks in predicting postoperative surgical complications: a retrospective study. Lancet Digit Health. 2021;3(8):e471–e85.

11. Dela Cruz CS, Wunderink RG, Christiani DC, Cormier SA, Crothers K, Doerschuk CM, et al. Future Research Directions in Pneumonia. NHLBI Working Group Report. Am J Respir Crit Care Med. 2018;198(2):256–63.

12. Bycroft C, Freeman C, Petkova D, Band G, Elliott LT, Sharp K, et al. The UK Biobank resource with deep phenotyping and genomic data. Nature. 2018;562(7726):203–9.

13. R Core Team (2020). R: A language and environment for statistical computing. R Foundation for Statistical Computing, Vienna, Austria. URL https://www.R-project.org/.

14. Pedregosa F, Varoquaux G, Gramfort A, Michel V, Thirion B, Grisel O, et al. Scikit-learn: Machine Learning in Python. J Mach Learn Res. 2011;12(null):2825–30.

15. Sigurdsson AI, Louloudis I, Banasik K, Westergaard D, Winther O, Lund O, et al. Deep integrative models for large-scale human genomics. Nucleic Acids Research. 2023.

16. Kingma D, Ba J. Adam: A Method for Stochastic Optimization. International Conference on Learning Representations. 2014.

17. Lundberg SM LS. A unified approach to interpreting model predictions. Advances in Neural Information Processing Systems 30 (NIPS). 2017.

18. Lawson EH, Louie R, Zingmond DS, Brook RH, Hall BL, Han L, et al. A comparison of clinical registry versus administrative claims data for reporting of 30-day surgical complications. Ann Surg. 2012;256(6):973–81.

19. Zhang T, Shang F, Ma Y, Xu Y, Sun W, Song H. Caveolin-1 Promotes the Imbalance of Th17/Treg in Chronic Obstructive Pulmonary Disease by Regulating Hsp70 Expression. Int J Chron Obstruct Pulmon Dis. 2023;18:565–74.

20. Williams TM, Lisanti MP. The Caveolin genes: from cell biology to medicine. Ann Med. 2004;36(8):584–95.

21. Jia W, Qi X, Li Q. Association Between Rs3807989 Polymorphism in Caveolin-1 (CAV1) Gene and Atrial Fibrillation: A Meta-Analysis. Med Sci Monit. 2016;22:3961–6.

22. Liu Y, Ni B, Lin Y, Chen XG, Chen M, Hu Z, et al. The rs3807989 G/A polymorphism in CAV1 is associated with the risk of atrial fibrillation in Chinese Han populations. Pacing Clin Electrophysiol. 2015;38(2):164–70.

23. Zhao L, Chen XG, Liu Y, Fang Z, Zhang F. Association of rs17042171 with chromosome 4q25 with atrial fibrillation in Chinese Han populations. Anatol J Cardiol. 2016;16(3):165–9.

24. Gudbjartsson DF, Arnar DO, Helgadottir A, Gretarsdottir S, Holm H, Sigurdsson A, et al. Variants conferring risk of atrial fibrillation on chromosome 4q25. Nature. 2007;448(7151):353–7.

25. L. Phan YJ, H. Zhang, W. Qiang, E. Shekhtman, D. Shao, D. Revoe, R. Villamarin, E. Ivanchenko, M. Kimura, Z. Y. Wang, L. Hao, N. Sharopova, M. Bihan, A. Sturcke, M. Lee, N. Popova, W. Wu, C. Bastiani, M. Ward, J. B. Holmes, V. Lyoshin, K. Kaur, E. Moyer, M. Feolo, and B. L. Kattman. ALFA: Allele Frequency Aggregator.” National Center for Biotechnology Information, U.S. National Library of Medicine. 2020.

26. Gao Y, Bertuccio CA, Balut CM, Watkins SC, Devor DC. Dynamin- and Rab5-dependent endocytosis of a Ca2+ -activated K+ channel, KCa2.3. PLoS One. 2012;7(8):e44150.

27. Yi SL, Liu XJ, Zhong JQ, Zhang Y. Role of caveolin-1 in atrial fibrillation as an anti-fibrotic signaling molecule in human atrial fibroblasts. PLoS One. 2014;9(1):e85144.

28. Ellinor PT, Lunetta KL, Glazer NL, Pfeufer A, Alonso A, Chung MK, et al. Common variants in KCNN3 are associated with lone atrial fibrillation. Nat Genet. 2010;42(3):240–4.

29. Dentali F, Sironi AP, Ageno W, Turato S, Bonfanti C, Frattini F, et al. Non-O blood type is the commonest genetic risk factor for VTE: results from a meta-analysis of the literature. Semin Thromb Hemost. 2012;38(5):535–48.

30. Stebbing J, Zhang H, Xu Y, Lit LC, Green AR, Grothey A, et al. KSR1 regulates BRCA1 degradation and inhibits breast cancer growth. Oncogene. 2015;34(16):2103–14.

31. Lee J, Seol MY, Jeong S, Kwon HJ, Lee CR, Ku CR, et al. KSR1 is coordinately regulated with Notch signaling and oxidative phosphorylation in thyroid cancer. J Mol Endocrinol. 2015;54(2):115–24.

32. Hung RJ, McKay JD, Gaborieau V, Boffetta P, Hashibe M, Zaridze D, et al. A susceptibility locus for lung cancer maps to nicotinic acetylcholine receptor subunit genes on 15q25. Nature. 2008;452(7187):633–7.

33. Schwartz AG, Cote ML, Wenzlaff AS, Land S, Amos CI. Racial differences in the association between SNPs on 15q25.1, smoking behavior, and risk of non-small cell lung cancer. J Thorac Oncol. 2009;4(10):1195–201.

34. Marwaha JS, Chen HW, Habashy K, Choi J, Spain DA, Brat GA. Appraising the Quality of Development and Reporting in Surgical Prediction Models. JAMA Surg. 2023;158(2):214–6.

